# Cost-Effectiveness of Systematic Screening and Treatment of Transthyretin Amyloid Cardiomyopathy (ATTR-CM) in Patients with Heart Failure with Preserved Ejection Fraction (HFpEF) in United States

**DOI:** 10.1101/2023.08.14.23294100

**Authors:** Anson TC Lau, Robert J. DiDomenico, Kibum Kim

**Author notes:** **Author’s Contribution** The conceptual idea was proposed in collaboration by all authors. All authors co-developed the cost-effectiveness model. Model analysis was performed by Anson Lau under the supervision of Kibum Kim and Robert DiDomenico. All authors reviewed the results and interpretation. Anson Lau and Kibum Kim compiled the draft manuscript. All co-authors reviewed, revised, and approved the manuscript. The overall research project was supervised by Kibum Kim, the corresponding author of this manuscript. **Correspondence** Kibum Kim, BPharm, MSc, PhD Address: 833 S Wood Street (m/c 871), Chicago, IL 60612. **Declaration of Interests** Anson Lau serves as a Senior Medical Advisor in AstraZeneca. Kibum Kim has received research funding from Renalytix AI, Grail LLC (a subsidiary of Illumina Inc.), and AstraZeneca. Robert DiDomenico has received research funding from the Chicago Department of Public Health, Cook County Department of Public Health, Otho S.A. Sprague Memorial Institute, and CSL Behring and has served on advisory boards for Abiomed and PhaseBio Pharmaceuticals.

## Abstract

**Background:** Transthyretin amyloid cardiomyopathy (ATTR-CM) is an underdiagnosed cause of heart failure in clinical practice. ^99m^Tc-pyrophosphate scintigraphy (PYP-scan) improves the accuracy of ATTR-CM detection, enabling timely initiation of tafamidis, a drug that slows the progression of ATTR-CM and lowers the risk of adverse cardiac events. We assessed the cost-effectiveness of universal systematic screening (USS) using PYP-scans in patients aged 60 years or older with heart failure with preserved ejection fraction (HFpEF) and ventricular wall thickness of at least 12mm.

**Methods:** Two screening strategies, USS of ATTR-CM with PYP-scan versus selected clinical referrals for PYP-scan in standard-of-care (SoC), were compared in a model-based assessment. Treatment decisions were based upon the accuracy of each screening strategy, which was followed by Markov state transitions across NYHA functional classes and death. Model inputs were identified from a literature review. We calculated lifetime cost in 2022 US dollars and quality adjusted life-years (QALYs) of each strategy. The primary outcome was the incremental cost-effectiveness ratio (ICER).

**Results:** The USS was associated with a significant increase in lifetime costs ($124,380 vs. $70,412) and modest improvement in QALYs (4.42 QALYs vs 4.36 QALYs). The ICER for the USS was $919,509 per QALY gained. ICER was sensitive to the age at the time of ATTR-CM diagnosis, true prevalence rate of ATTR-CM, and daily cost of tafamidis.

**Conclusions:** Owing to the high cost of treatment with tafamidis, USS along with PYP scan for ATTR-CM in older HFpEF patients with ventricular wall thickening is unlikely to become a cost-effective strategy at a liberal WTP threshold.

**What is Known:**

- ATTR-CM is a rare but serious condition that leads to a restrictive cardiomyopathy with symptoms that mimic heart failure from other causes (e.g., HFpEF).
- PYP-scan, when it is used along with the other conventional diagnostic tests, is a non-invasive diagnostic tool that sensitively and specifically detect ATTR-CM followed by a timely treatment with tafamidis.
- Tafamidis can slow down ATTR-CM progression and improve survival.

**What the Study Adds:**

- This is the first attempt to evaluate the cost-effectiveness of universal systematic screening of ATTR-CM using PYP-scans in older HFpEF patients with ventricular wall thickening.
- The study found that the universal systematic screening is not cost-effective compared to standard-of-care at a liberal WTP threshold, mainly due to the costly treatment option.
- The study identified the key parameters that affect the cost-effectiveness of screening, such as age, prevalence, and drug cost.

## Introduction

Transthyretin amyloid cardiomyopathy (ATTR-CM) is a rare, progressive disease characterized by the buildup of misfolded transthyretin (TTR) forming amyloid fibrils in the myocardium.^1^ The pathophysiological progression of ATTR-CM causes a restrictive cardiomyopathy and symptoms of heart failure. Once patients with ATTR-CM become symptomatic, their quality of life (QoL) rapidly deteriorates.^1^ However, because of the clinical manifestation is similar to the other types of heart failure, ATTR-CM is one of the most underdiagnosed heart diseases in the US healthcare system.^2^ The poor disease awareness and suboptimal sensitivity of the routine clinical assessment have been pointed out as the main reasons for the undiagnosed ATTR-CM cases. ^2^

Tafamidis, the first-in-class disease-modifying transthyretin (TTR) kinetic stabilizer, has been approved for treatment of adult patients with ATTR-CM since May 2019.^3^ Compared to the standard of care (SoC), tafamidis delayed myocardial amyloid progression^4^, significantly reduced all-cause mortality (hazard ratio [HR] = 0.59, 95% confidence interval [CI] 0.44– 0.79, P<0.001)^5^, decreased CV-related hospitalizations (relative risk [RR] = 0.68, 95% CI 0.56–0.81)^6^, and improved health-related QoL.^7^ Thus, timely diagnosis and treatment with tafamidis is expected to improve overall QoL and longevity.^8^ Despite the anticipated clinical effectiveness, the use of tafamidis is highly respective and contingent upon the completion of extensive ATTR-CM screening, due to the acquisition cost exceeding $200,000 annually.^9^

^99m^>Tc-pyrophosphate bone scintigraphy (PYP-scan) is a non-invasive nuclear imaging technique that enables detection of cardiac amyloidosis. In combination with serum free light-chain (FLC) test or immunofixation electrophoresis (IFE) to rule out light chain amyloidosis, the PYP-scan achieved an overall sensitivity of 92.2% in detecting cardiac amyloidosis among patients with underlying ATTR-CM.^10^ Despite the accuracy, utilization of the PYP-scan in clinical practice remains low, preventing healthcare systems from expedited diagnosis of ATTR-CM.^11^ Currently, PYP-scans are used selectively to screen high-risk patients with increased ventricular wall thickness and clinical sequelae suggestive of amyloidosis (e.g. carpel tunnel syndrome, polyneuropathy, atrioventricular block).^2,12,13^ A recent community-based screening project using universal PYP-scans in patients aged 60 years or older with heart failure with preserved ejection fraction (HFpEF) and ventricular wall thickness of at least 12mm resulted in a higher rate of ATTR-CM diagnosis compared to standard clinical diagnosis (6.3% vs 1.3%).^14^ The difference in the detection rates suggests that a systemic screening, once universally administered, potentially close the diagnostic gap in real-world settings.

The decision to adopt a new healthcare technology as part of the SoC should be further informed by a value assessment, including the long-term clinical benefits and cost associated with the treatment decision. The purpose of this study was to determine the long-term cost-effectiveness of universal systematic screening (USS) using PYP-scans to identify ATTR-CM in older patients with HFpEF and ventricular wall thickening compared to the SoC in which selective screening is used based on clinical suspicion.

## Methods

### Overview, target population and model structure

A model-based cost-effectiveness assessment was performed under the US healthcare system perspective. We developed a *de novo* model to compare the lifetime healthcare costs and quality-adjusted life years (QALYs) of the two screening strategies, USS and SoC. Study population includes patients with HFpEF and ventricular wall thickness of at least 12 mm. Our model utilized a cohort of patients aged 60 years or older with a median age of 78 years where the estimated true ATTR-CM prevalence of 6.8%.^15^ The proportion of patients at the NYHA I – IV was 10.4%, 34.7%, 33.1%, 21.9%.^8,14,16,17^

In the USS group, all patients with HFpEF and ventricular wall thickening undergo a one-time PYP–scan, in combination with FLC testing or IFE, to determine ATTR-CM and exclude light chain amyloidosis. In the SoC group, selected patients with HFpEF (12.2%) undergo a PYP-scan to screen for ATTR-CM based on clinical suspicion.^15^ (Figure 1) For any positive ATTR-CM diagnosis, a TTR genetic test will be used to confirm the mutation type. The hypothetical cohort enters the decision model through either the USS or SoC path. A diagnosis of ATTR-CM would be made upon the accuracy of each screening strategy, which is followed by treatment with tafamidis.

**Figure 1:**
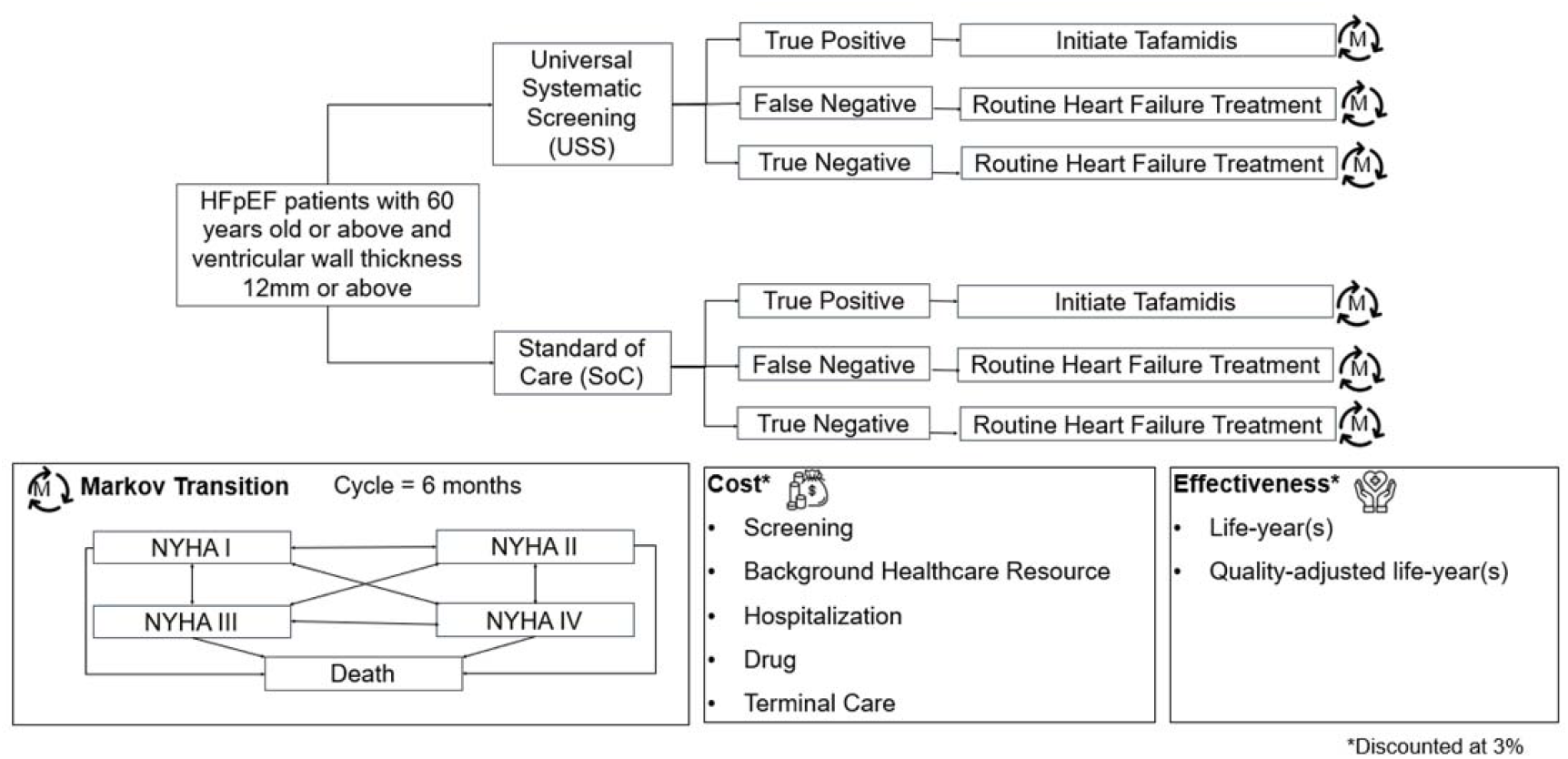
Model Structure.

The chance nodes describing treatment choice upon the diagnostic accuracy were followed by Markov state transitions simulating disease progression over the life-time horizon. Health states included New York Heart Association (NYHA) Function Classes I, II, III, and IV; the terminal state is death. The model cycle was 6 months and outcomes were calculated over the lifetime horizon. The model progresses on a six-month cycle. We applied an annual discount rate of 3% for both cost and QALY. The model was developed using TreeAge Pro (Healthcare Version) 2022 (TreeAge Software Inc., Williamstown, MA).

### Model Inputs - Screening performance and treatment scenarios

USS performs better than SoC in identifying ATTR-CM with an improved sensitivity without losing specificity.^14^ Based on the previous literature review, we assumed diagnostic sensitivities of 92.2% and 19.0% for USS and SoC, respectively.^10,14^ Combining the prevalence of ATTR-CM and testing performance, the proportion of HFpEF patients diagnosed with ATTR-CM using USS was 6.3% compared to 1.3% using the SoC.^14^ Both USS and SoC were assumed to be 100% specific in excluding patients without underlying ATTR-CM. Thus, the target population followed one out of the three clinical scenarios: true-positive ATTR-CM cases treated with tafamidis, false-negative ATTR-CM patients treated without tafamidis, and true-negative cases without ATTR-CM. (FIGURE 1)

### NYHA-Class Progression and Treatment Effectiveness

The transition probabilities across NYHA classes I though IV were extracted from the Public Summary Document of Tafamidis in Australia Pharmaceutical Benefits Scheme (PBS).^18^ We applied the transition rate ratio across the progressions, to calculate the Markov state-specific transition probabilities.^19^ (Supplement) Tafamidis treatment lowered risk of cardiovascular (CV) hospitalization.^20^ We abstracted the CV hospitalization rate from published data and adjusted for potential immortal bias using the survivor average causal effect (SACE) approach to calculate the model inputs.^20,21^

Mortality is a function of age, NYHA class, ATTR-CM and tafamidis treatment. We used the age-specific mortality rate from the United States in Human Mortality Database 2020 as baseline input and applied the HR of death for HF patients versus general population.^21,22^ HF-specific mortality was further adjusted by the rate ratio of death across the NYHA classes and between ATTR-CM versus non-ATTR-CM HF to calculate the MArkove state-specific mortality.^23,24^ To account for the mortality benefits of tafamidis, we further applied the HR of death for tafamidis versus placebo obtained from for the ATTR-ACT study.^5^

### Costs and Utilities

Screening and intervention costs were defined by the review of the 2021 Center for Medicare and Medicaid Services (CMS) Physician fee schedule.^25^ We determined the PYP-scan cost input by aggregating the physician and facility charges for the HCPCS procedure code (A9538).^25^ The cost of the FLC test, IFE and TTR genetic test were directly abstracted from the publicly available laboratory service charge tables.^26–28^

The differences between the USS and SoC groups in correctly identifying and treating patients will influence healthcare costs and healthcare resources utilization (HRU), including hospital admissions and visits to primary care, specialty care, and the emergency department (ED). We estimated the NYHA class-specific per-cycle HRU using the Canadian Agency for Drugs & Technologies in Health (CADTH) data.^29^ To reflect the cost from the US perspective, the expected annual resource use was multiplied by the unit cost of service available from the Agency for Healthcare Research and Quality.^30–32^ All the costs inputs were adjusted to the March 2022 value in USD using Consumer Price Index for All Urban Consumers: Medical Care in U.S. City Average.^33^

Patients would initiate and continue to receive tafamidis upon the diagnosis of ATTR-CM. Aside from tafamidis therapy, all patients would continue being treated with other therapies to treat heart failure (e.g., angiotensin converting enzyme inhibitors, beta-blockers, diuretics, antiplatelets, and anticoagulants).^14^ The cost of tafamidis and other baseline medications were extracted from RedBook wholesale acquisition price.^9^.

QALY was a function of health utility score for each NYHA class and longevity. We extracted utility scores from the ATTR-ACT trial and long-term extension study.^34^ Health utility decreased with the NYHA class progressions from 0.893 (class I) to 0.406 (class IV) when patients received placebo for ATTR-CM. Our model reflected the regimen- and NYHA class-specific utility scores to calculate the QALY gains. In addition, our model accounted for the negative impact of hospitalizations on QoL, which was multiplied by the anticipated length of hospital stay per cycle.^19^

### Analysis

The primary outcome measure was the incremental cost-effectiveness ratio (ICER), defined as incremental costs per QALY gained for USS versus SoC over the lifetime. We compared the ICER calculation with the stringent and liberal willingness-to-pay (WTP) thresholds, $100,000 and $200,000. Both lifetime and 5-year short-term outcomes were reported.

For one-way sensitivity analyses. we used the 95% CI range of each input. When the CI or standard error range was not available, ± 10% of the base-case input was used. We anticipated that the cost of tafamidis and screening performance (i.e., sensitivity) would be the primary interests of stakeholders. Therefore, we evaluated the impact of both inputs simultaneously by performing two-way sensitivity testing. Also we calculated ICER against changes in the age to assess the impact of diagnosis made in different ages.

We simulated an extreme case in which the best clinical scenario was modeled using early (60 years of age) detection of ATTR-CM via USS, all of whom were NYHA class I at the time of diagnosis and treated with tafamidis once diagnosed. We also performed a threshold analysis, calculating the maximum affordable tafamidis price at various WTP thresholds.

## Results

### Base-Case Results

USS was associated with an increase in both cost and QALYs. The lifetime cost and effectiveness for the USS strategy were $124,380 and 4.41 QALYs, respectively, over the median survival of 7.5 years. The SoC strategy incurred a lifetime cost of $70,412 and yielded 4.36 QALYs over a median survival of 7.0 years. Thus, the lifetime ICER was $919,509/QALY gained. (Table 2). When we changed the time horizon of the model from lifetime to five years, the ICER increased to $1,223,719/QALY gained (incremental cost: $44,698; incremental effectiveness: 0.04 QALYs gained).

**Table 1:** Selected Estimates of Parameters Used in the Model.

| Variable | Value | Range of SA | Reference |
| --- | --- | --- | --- |
| **Proportion of patients receiving PYP-scan** |  |  |  |
| USS | 100% | N/A |  |
| SoC | 12.2% | 11.0% – 13.4% | 15 |
| **Age of Patients** |  |  | 14 |
| Median age of ATTR-CM diagnosis | 84 | 70 – 98 |  |
| Median age of normal HF phenotype | 78 | 64 – 92 |  |
| **Baseline NYHA Class Distribution in Diagnosed ATTR-CM Patients** |  |  | 14,16 |
| ATTR-CM, true positive, NYHA I | 0.104 | 0.094 – 0.114 |  |
| ATTR-CM, true positive, NYHA II | 0.347 | 0.312 – 0.381 |  |
| ATTR-CM, true positive, NYHA III | 0.331 | 0.298 – 0.364 |  |
| ATTR-CM, true positive, NYHA IV | 0.219 | - |  |
| **Prevalence** |  |  | 14,43 |
| True Prevalence of ATTR-CM | 0.068 | 0.041 – 0.106 |  |
| Detected Prevalence of ATTR-CM in USS | 0.063 | 0.038 – 0.098 |  |
| Detected Prevalence of ATTR-CM in SoC | 0.013 | 0.007 – 0.021 |  |
| **Performance of Screening Strategy** |  |  | 43 |
| USS: Sensitivity | 0.922 | 0.89 – 0.95 |  |
| SoC: Sensitivity | 0.190 | 0.102 – 0.307 |  |
| USS or SoC: Specificity | 1 |  |  |
| **Cost** |  |  |  |
| **Cost of Screening** |  |  |  |
| Cost of PYP-scan | \$4263 | \$3836 – \$4689 | 25 |
| Cost of kappa free light chain test (FLC) | \$439 | \$395 – \$483 | 26 |
| Cost serum immunofixation electrophoresis (IFE) | \$119 | \$108 – \$131 | 27 |
| Cost of Transthyretin Amyloidosis Genetic Test | \$500 | \$450 – \$550 | 28 |
| **Cost of 6-month Background Healthcare Resource Use** |  |  |  |
| Cost of 6-month resource in NYHA I in ATTR-CM | \$514 | \$462 – \$565 | 29-31 |
| Cost of 6-month resource in NYHA II ATTR-CM | \$1327 | \$1195 – \$1460 | |
| Cost of 6-month resource in NYHA III ATTR-CM | \$2564 | \$2308 – \$2820 | |
| Cost of 6-month resource in NYHA IV ATTR-CM | \$4840 | \$4356 – \$5324 | |
| **Cost of Hospitalization** |  |  |  |
| Cost of hospitalization per day | \$4025 | \$3623 – \$4428 | 32 |
| **Cost of 6-month Drug Treatment** |  |  |  |
| Cost of one tafamidis 61 mg capsule | \$652.5 | \$587 – \$718 | 9 |
| Cost of 6-month drug treatment (with tafamidis) in true-positive ATTR-CM | \$121598 | \$109438 – \$133758 | 14,9 |
| Cost of 6-month drug treatment (without tafamidis) in false-negative ATTR-CM | \$2517 | \$2265 – \$2768 | |
| **Cost of terminal care per mortality event** | \$9148 | \$8233 – \$10623 | 44 |
| **Effectiveness** |  |  |  |
| **Cardiovascular Hospitalization Rate in 6-month** |  |  | 20,45 |
| ATTR-CM, NYHA I/II; Tafamidis | 0.169 | 0.143 – 0.200 |  |
| ATTR-CM, NYHA I/II; Placebo | 0.355 | 0.304 – 0.414 |  |
| ATTR-CM, NYHA III/IV; Tafamidis | 0.262 | 0.241 – 0.284 |  |
| ATTR-CM, NYHA III/IV; Placebo | 0.345 | 0.317 – 0.374 |  |
| **Length of Stay of Cardiovascular Hospitalization** |  |  | 20 |
| ATTR-CM, NYHA I/II; Tafamidis | 8.50 | 7.12 – 9.89 |  |
| ATTR-CM, NYHA I/II; Placebo | 9.64 | 8.15 – 11.1 |  |
| ATTR-CM, NYHA III/IV; Tafamidis | 8.91 | 8.65 – 9.18 |  |
| ATTR-CM, NYHA III/IV; Placebo | 9.42 | 8.80 – 10.0 |  |
| **Hazard Ratio of Mortality** |  |  | 21-24,5 |
| Healthy (Reference) | 1.0 |  |  |
| ATTR-CM, NYHA I, Tafamidis | 1.45 | 1.30 – 1.59 |  |
| ATTR-CM, NYHA II, Tafamidis | 2.57 | 2.32 – 2.83 |  |
| ATTR-CM, NYHA III, Tafamidis | 5.90 | 5.31 – 6.50 |  |
| ATTR-CM, NYHA IV, Tafamidis | 9.64 | 8.68 – 10.6 |  |
| ATTR-CM, NYHA I, Placebo | 2.59 | 2.33 – 2.85 |  |
| ATTR-CM, NYHA II, Placebo | 4.59 | 4.13 – 5.05 |  |
| ATTR-CM, NYHA III, Placebo | 9.08 | 8.17 – 9.98 |  |
| ATTR-CM, NYHA IV, Placebo | 14.8 | 13.3 – 16.3 |  |
| **Utility** |  |  | 34 |
| ATTR-CM, NYHA I, Tafamidis | 0.874 | 0.854 – 0.994 |  |
| ATTR-CM, NYHA II, Tafamidis | 0.832 | 0.812 – 0.852 |  |
| ATTR-CM, NYHA III, Tafamidis | 0.707 | 0.687 – 0.727 |  |
| ATTR-CM, NYHA IV, Tafamidis | 0.558 | 0.440 – 0.596 |  |
| ATTR-CM, NYHA I, Placebo | 0.893 | 0.873 – 0.932 |  |
| ATTR-CM, NYHA II, Placebo | 0.802 | 0.782 – 0.822 |  |
| ATTR-CM, NYHA III, Placebo | 0.706 | 0.686 – 0.726 |  |
| ATTR-CM, NYHA IV, Placebo | 0.406 | 0.289 – 0.477 |  |
| Hospitalization, NYHA I | -0.04 | - | 19 |
| Hospitalization, NYHA II | -0.07 | - |  |
| Hospitalization, NYHA III | -0.1 | - |  |
| Hospitalization, NYHA IV | -0.29 | - |  |

**Table 2:**
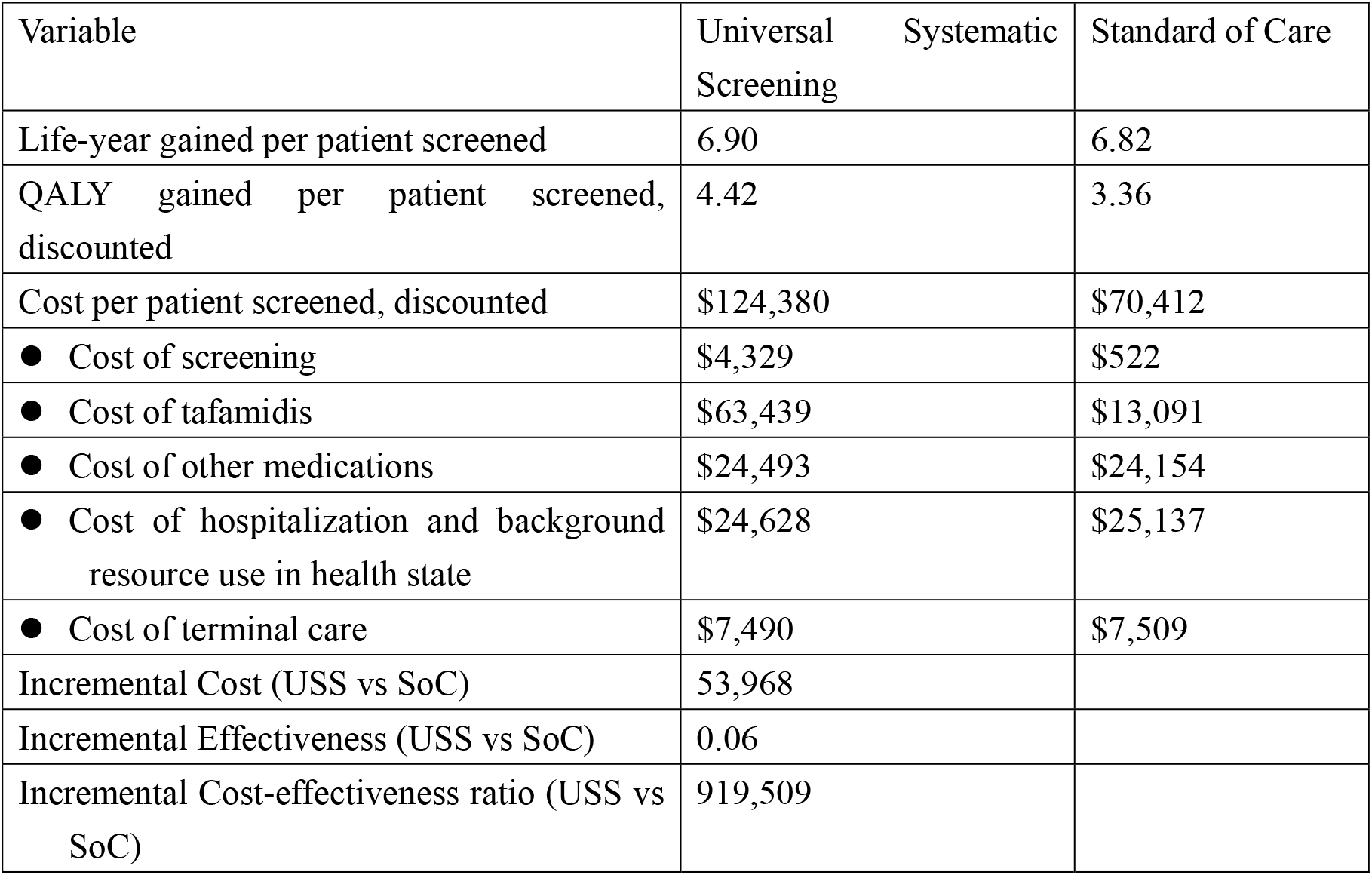
Base-Case Cost-effectiveness Results.

**Table 3:** Price threshold analysis of tafamidis.

| WTP | Threshold | Percent change from original price (\$652.5) |
| --- | --- | --- |
| 100000 | \$29 | - 96% |
| 150000 | \$67 | - 90% |
| 200000 | \$105 | - 84% |

### Sensitivity Analyses

From the one-way sensitivity analyses, ICER varied most in response to changes in age at the time of ATTR-CM diagnosis and daily cost of tafamidis. (Figure 2) Other influential factors included the true prevalence of ATTR-CM, sensitivity of identifying ATTR-CM using the USS strategy, HR of ATTR-CM mortality for tafamidis versus placebo. Health-related utility of each NYHA classes is also an influential input. (Figure 2) Our threshold analysis demonstrated that the daily cost of tafamidis would have to be less than $105, 84% lower than the current wholesale acquisition price, to yield an ICER less than the liberal WTP threshold of $200,000/QALY. When we calculated ICER on various age at the current wholesale acquisition price, the ICER decreased as much as 26% when ATTR-CM was diagnosed at older ages but increased slightly with increasing age at lower tafamidis acquisition costs necessary to meet the WTP thresholds. (Figure 3). In the scenario where tafamidis would achieve maximum clinical benefits, the ICER estimate became $1,139,632 per QALY; the USS strategy gained an additional 0.08 QALYs with additional cost of $92,293 per patient screened.

**Figure 2:**
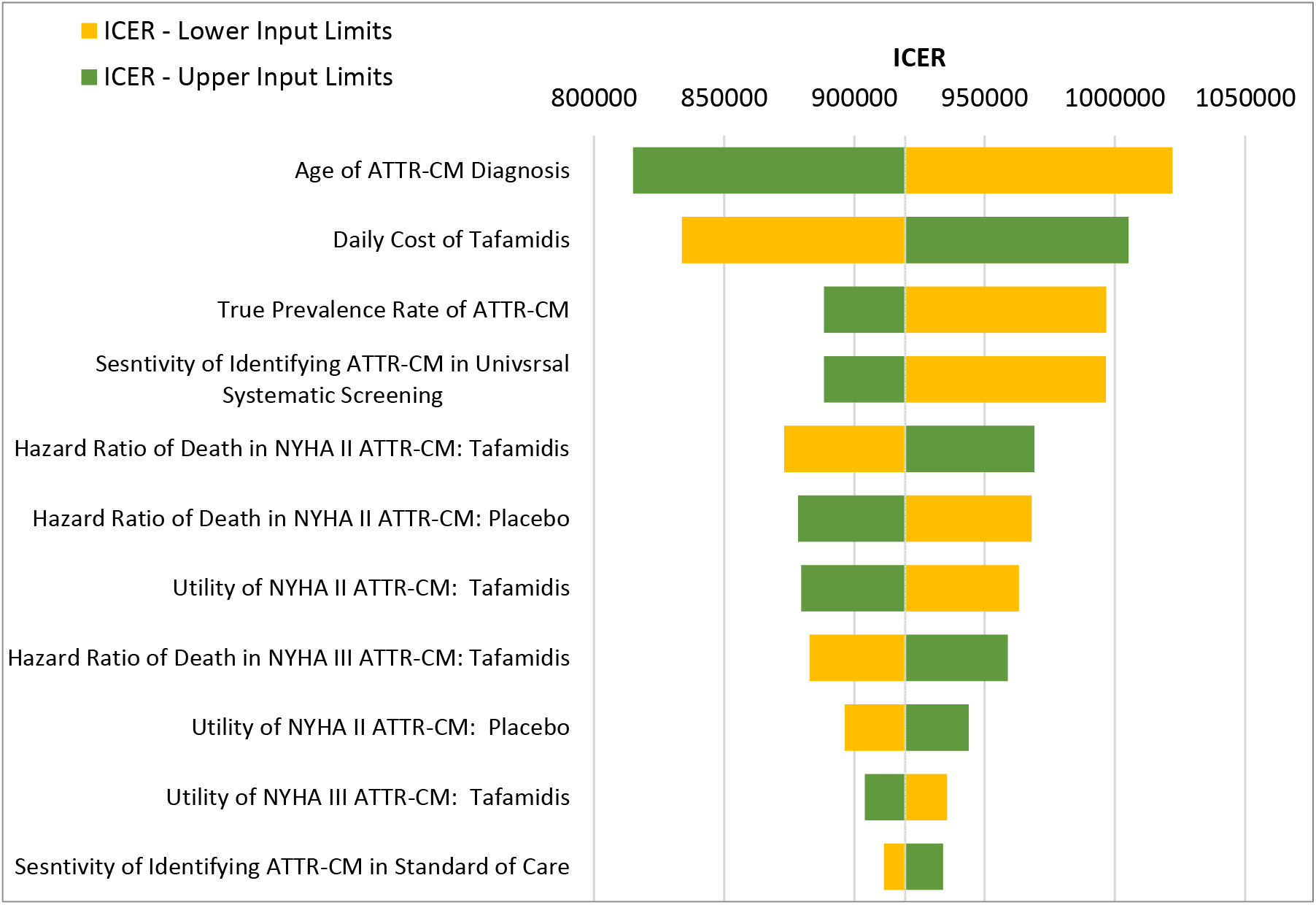
Tornado diagram of 1-way Sensitivity Analysis.

**Figure 3:**
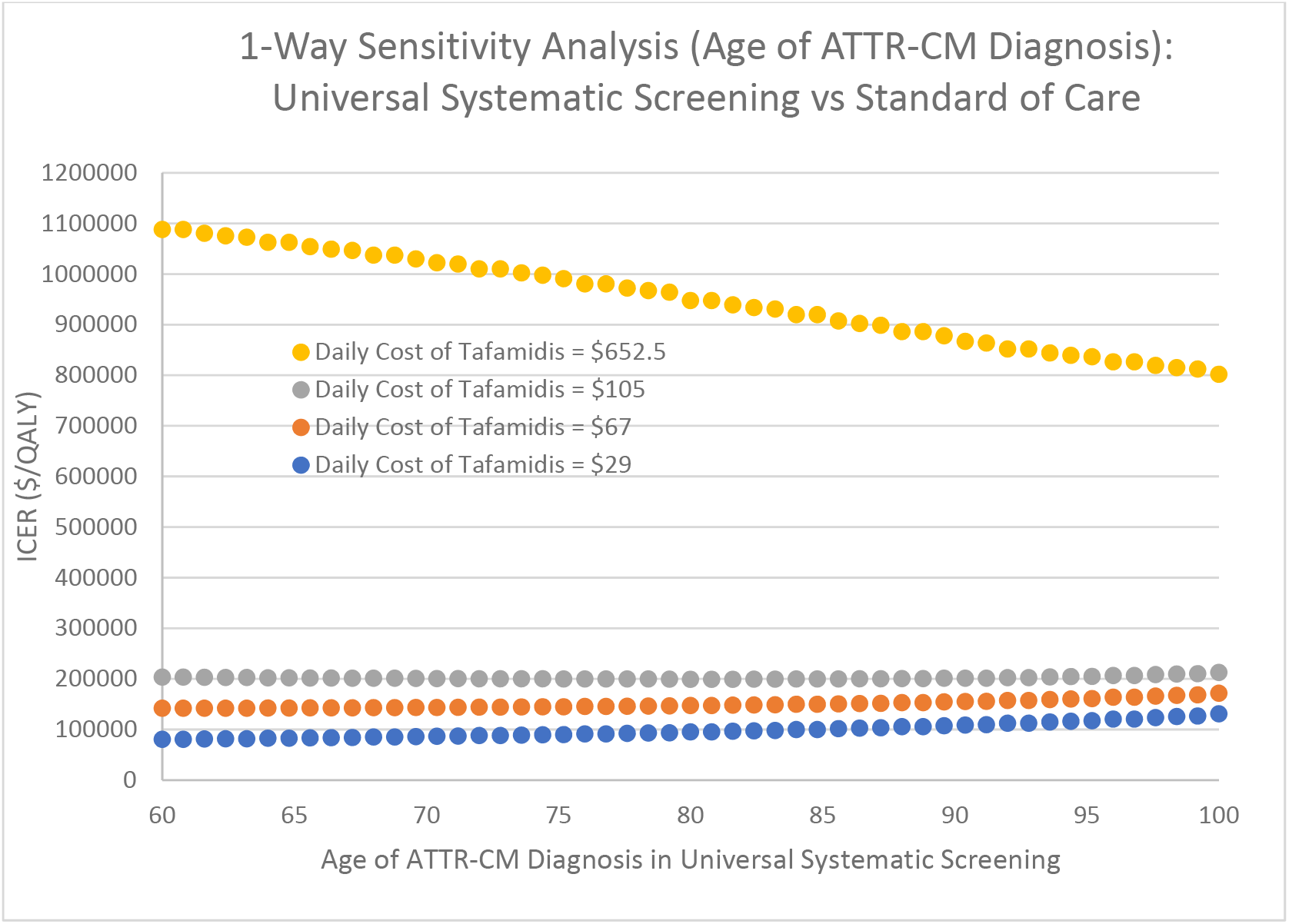
1-Way Sensitivity Analysis: Age of ATTR-CM Diagnosis with USS at Different Cost of Tafamidis.

## Discussion

Our study demonstrated that a USS strategy to identify ATTR-CM in patients with HFpEF and ventricular wall thickening followed by tafamidis use was effective but very costly approach options. The ICER of USS-guided tafamidis treatment ($919,509/QALY) is higher than the four-fold of the generally acceptable, liberal WPT threshold. The cost-effectiveness of a USS strategy for detecting and treating ATTR-CM was most sensitive to age at the time of ATTR-CM diagnosis, cost of tafamidis, true prevalence of ATTR-CM among patients with HFpEF, and the sensitivity of the USS strategy detecting ATTR-CM. Nevertheless, the conclusion was robust using reasonable assumptions for included variables.

Our *de-novo* model is structurally distinctive from previous cost-effectiveness studies in ATTR-CM. An economic outcome assessment by Kazi et al. in patients with confirmed ATTR-CM projected an ICER of $880,000 for tafamidis compared to a non-specific treatment approach.^35^ While Kazi’s model reflected an inexorably progressive mortality inputs and projected a fairly accurate life-year estimate, similar to the clinical trial output,^35,36^ it did not address the health utility changes associated with a decline in functional capacity as we did in our model.^35,37^ Therefore, our approach would improve the accuracy in estimating the incremental QALY gained. Another cost-effectiveness model by Ge et al. compared several screening strategies to detect ATTR-CM but did not incorporate the QoL change against the ATTR-CM progression that we incorporated into our model.^39^ Compared to the previous studies, we modeled more comprehensive factors that influence treatment decisions and outcomes, including screening accuracy, cost, utility, rate of cardiovascular complication for each of the NYHA functional classes, and mortality. We improved a face validity by reflecting the high-level of granularity in assessing outcomes.

For example, the respective life-time effectiveness and cost estimates for the Ge’s study population were 7.2 to 7.3 QALYs and approximately $210,000 treatment cost, while we calculated around 4.4 QALYs slightly over the 7-year life expectancy. We suspect the difference in the effectiveness estimates were attributed to several factors. Our approach addressed the disutility of ATTR-CM progression toward the advanced NYHA classes, which further diminish the anticipated lifetime QALYs in the elderly population. Further, our model reflects stage-specific hospitalization costs^32^ and recent acquisition costs of medications. ^9^ Such updates, besides the model structural distinct from the previous model-based assessment calculated the ICER higher than the previous economic outcome estimates.

Although early diagnosis and timely tafamidis initiation using USS is expected to improve survival and QoL for ATTR-CM patients^8^, it does not appear to be a cost-effective approach for healthcare systems. In particular, the high acquisition cost may limit access to and uptake of this disease-modifying treatment.^35^ The cost-effectiveness of widespread implementation of USS for ATTR-CM may be limited unless there is a dramatic decrease in the cost of tafamidis, which was $652.5 daily ($238,163 annually) in 2022.^9^ We estimated that a USS strategy to detect ATTR-CM would be a cost-effective approach if the tafamidis acquisition cost decreased by 84 – 96 % (at WTP thresholds of $200,000/QALY and $100,000/QALY, respectively). Our conclusion is generally consistent with previous analyses that suggested a 93% price reduction would be required for tafamidis to be a cost-effective treatment for ATTR-CM at a WTP threshold of $100,000/QALY.^35,38^

At the current acquisition cost of tafamidis, we estimated that the ICER increases with the earlier diagnosis of ATTR=CM within the younger population. Using significantly lower tafamidis acquisition costs necessary to achieve cost-effectiveness at accepted WTP thresholds, the influence of early diagnosis and treatment of ATTR-CM on ICER values was less. The scenario analysis highlights the inverse relationship between age at the time of ATTR-CM diagnosis and ICER, resulting in an even higher ICER ($1,139,632/QALY), primarily driven by the synergy of excessive tafamidis cost and longer survival. Together with prior analyses, our study highlights the need for drug pricing mechanisms that lower overall drug costs and improve the cost-effectiveness of tafamidis, such as value-based pricing and outcome-based contracting^39^, that may remove cost as a barrier and improve patient access to advanced screening and treatment strategies.

Despite increasing ATTR-CM detection by 6-fold, the USS strategy we modeled was associated with a low positivity rate (6.3%), limiting its cost-effectiveness. Additional selection criteria, such as the ATTR-CM score, may help guide use of ^99m^Tc-PYP, enhance screening success in selected high-risk HFpEF patients, and improve the cost-effectiveness of this approach.^40^ The ATTR-CM score incorporates 3 clinical (age, male sex, hypertension diagnosis) and 3 echocardiographic (ejection fraction, posterior wall thickness, relative wall thickness) variables to identify ATTR-CM in HFpEF cohorts. An ATTR-CM score ≥6 has a positive predictive value ≥25% resulting in an ATTR-CM prevalence of ≥10% among HFpEF patients. ^40^ The advancement of artificial intelligence and machine learning can be considered to improve the diagnosis performance and may lower the ICER for USS further.^41,42^

## Limitation

Our model has several limitations that must be considered. First, our model inputs, including Markov transition probabilities, NYHA distribution, and mortality, are derived from multiple sources due to limited data availability. Some input sources are from European countries and may not accurately represent the US practices. We estimated the baseline distribution of NYHA class based on the median age of ATTR-CM diagnosis, using data from Germany in which the severity of ATTR-CM may differ compared to the normal phenotype in the US at the same age. Second, drug adherence was assumed to be consistent in the model. In the real-world, drug adherence may be limited by financial barriers, patient comorbidities, polypharmacy, and side effects that may affect populations differently, resulting in inconsistent adherence. Besides the specific limitation of our approach, general limitations of a Markov model-based cost-effectiveness analysis owing to the lack of memory effect and standardized clinical scenario should be considered.

## Conclusion

USS to detect ATTR-CM in patients ≥ 60 years with HFpEF and ventricular wall thickening is unlikely to become a cost-effective strategy unless the cost of tafamidis is substantially lowered. The cost-effectiveness of a USS approach to detect ATTR-CM is also sensitive to the changes in several factors including the age at the time of ATTR-CM diagnosis, true prevalence of ATTR-CM among patients with HFpEF, and the sensitivity of the USS approach for detecting ATTR-CM. The high ICER is mainly driven by the drug cost. Despite the advancement of non-invasive imaging techniques and disease-modifying treatments, further utilization of the systematic screening should be accompanied with affordable treatment options.

## Data Availability

All the completed analysis results are included in the manuscript. Should the additional report be generated, additional data will be available upon the request from editorial offices.

